# Raman spectroscopy, used transcutaneously and non-invasively from a finger, to predict COVID-19: A feasibility, proof-of-concept study

**DOI:** 10.1101/2023.01.19.23284747

**Authors:** Allen B. Chefitz, Thomas Birch, Yongwu Yang, Arib Hussain

## Abstract

**BACKGROUND:** A definitive COVID-19 infection typically is diagnosed by laboratory tests, including real-time, reverse-transcriptase Polymerase Chain Reaction (PCR)-based testing. These currently available COVID-19 tests require the patient to provide an extra-corporeal specimen and the results may not be immediate. Consequently, a variety of rapid antigen tests for COVID-19, all with a wide range of accuracy in terms of sensitivity and specificity, has proliferated (1,2). These rapid tests now represent a significantly larger proportion of all testing done for COVID-19, yet suffer from requiring a physical specimen from the nose or mouth and waiting 15 minutes for most.

As a solution, we propose a non-invasive, trans-cutaneous, real-time viral detection device, based on the principles of Raman spectroscopy and machine learning. It does not require any extra-corporeal specimens and can be configured for self-administration. It can be easily used by non-experts and does not require medical training. Our approach suggests that our non-invasive, transcutaneous method may be broadly useful not only in COVID-19 diagnosis, but also in other diagnoses.

**METHODS:** 160 COVID positive (+) patients and 316 COVID negative (-) patients prospectively underwent nasal PCR testing concurrently with testing using our non-invasive, transcutaneous, immediate viral detector. Both the PCR and our experimental viral detector tests were performed side-by-side on outpatients (N=389) as well as inpatients (N= 87) at Holy Name Medical Center in Teaneck, NJ between June 2021 and August, 2022. The spectroscopic data were generated using an 830nm Raman System with SpectraSoft (W2 Innovations)and then, using machine learning, processed to provide an immediate prediction. A unique patient-interface for finger insertion enabled the application of Raman spectroscopy to viral detection in humans.

**RESULTS:** The data analysis algorithm demonstrates that there is an informative Raman spectrum output from the device, and that individual Raman peaks vary between cases and controls. Our proof-of-concept study yields encouraging results, with a specificity for COVID-19 of 0.75, and a sensitivity (including asymptomatic patients) of 0.80.

**CONCLUSIONS:** The combination of Raman spectroscopy, artificial intelligence, and our unique patient-interface admitting only a patient finger achieved test results of 0.75 specificity and 0.80 sensitivity for COVID-19 testing in this first in human proof-of-concept study. More significantly, the predictability improved with increasing data.

## Introduction

The gold standard for establishing the diagnosis of infection with SARS-CoV-2, otherwise known as COVID-19, is the real time, reverse transcriptase polymerase chain reaction (PCR) test. Another diagnostic test, less specific and sensitive, is the lateral flow assay. Currently available PCR and lateral flow assay tests require a specimen, such as sputum, saliva, urine, stool, or breath. We formulated a hypothesis that COVID-19 could be detected transcutaneously in a finger in a way similar to pulse oximetry measurements. Raman spectroscopy, a spectroscopic technique that determines vibrational modes of molecules, has been used to detect the unique spectrum of SARS-CoV-2, as well as other viruses and molecules, using an extra-corporeal specimen (see Fig.1) (3,4,5). Carlomagno et al (6) describe a Raman-based classification model to discriminate the signal in saliva of COVID-19 patients. Peaks located at 748, 922, 1048, 1249, and 1126 presented “significant differences from a preliminary visual inspection” between the different experimental groups. Peaks at 1048 and 1126 cm-1 are believed to be attributable to the tryptophan and phenylalanine signal and to the C-N and C-C stretching. Aromatic amino acids, including tryptophan, play an important role in the spike protein structure. However, Raman detecting COVID-19 non-invasively, transcutaneously from the blood has not been established. The Raman Shift that SARS-CoV-2 produces in a saliva, sputum, or urine specimen may not be the same shift as seen in the blood using spectroscopy non-invasively with a transcutaneous detection system. Further, although a database currently exists for Raman spectra of SARS-CoV-2 and many other viruses and molecules, those databases are with extra-corporeal specimens placed on a platform. No such database exists for blood-borne SARS-CoV-2 when viewed transcutaneously. We set out to target not only SARS-CoV-2, likely at a low concentration, but also the entire mixture of blood components that is typically seen in COVID-19.

**Figure 1:**
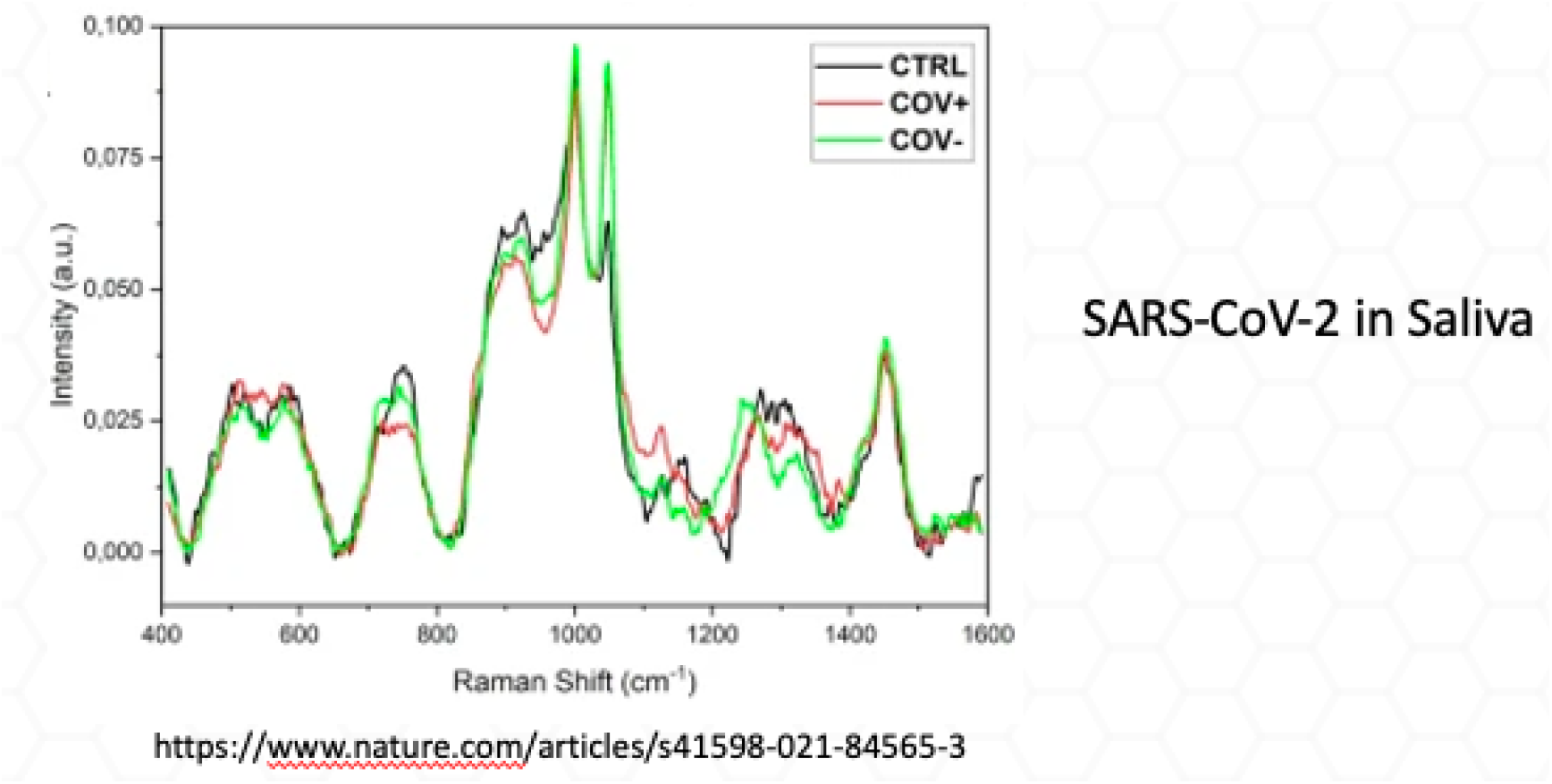

Despite a high signal to noise ratio (SNR) of our device, 6400:1, we hypothesized that unless the concentration of SARS-CoV-2 in the bloodstream were high enough, Raman would perhaps fail to detect the actual SARS-CoV-2 RNA in the blood transcutaneously or even directly. In designing our study of Raman applied to a finger, we considered testing our hypothesis using blood samples in a tube or container prior to the step of applying the laser to a finger. However, the process of removing blood and storing it ex-vivo would alter the blood components in a way that would significantly change our in-vivo transcutaneous and non-invasive analysis of what is in the blood unique to COVID-19. Further, several reports (7,8) of the systemic effects and sequelae of COVID-19, involving the heart, liver, clotting system, inflammatory system, intestines, kidney, and brain, convinced us that there must be several molecular targets in the blood of COVID-19 patients. Ivayla Roberts et al describe “untargeted metabolomics of COVID-19 patient serum…” and add to previous studies reporting metabolomics in the blood. They present a “high dimensional predictive model…, based on more than 900 metabolites. Next with the goal of clinical application in mind…present results from a predictive model based on 20 compounds selected largely on the basis of our confidence in the metabolites’ identity and known biological function.” They also find that “deoxycytidine and kynurenine remained significant regardless of patient demographics or underlying conditions’’ (9). They also cite studies “showing that the major plasma metabolic changes were fatty acids in men and glycerophosphocholines and carbohydrates in women (9). Given all the evidence for biomarkers in the blood in patients with COVID-19, we hypothesized that we could target the unique combinations and concentrations of multiple blood-borne molecules, in addition to SARS-CoV-2 RNA, using Raman transcutaneously. We would combine the optical engineering of Raman (detecting the signals) with machine learning (classifying and predicting) to arrive at a diagnosis of COVID-19.

Balan et al (28) review spectroscopic methods as applied “from molecular to clinical practice” and explain the advantages of using Raman spectroscopy. In discussing infrared spectroscopy-near-infrared, mid-infrared, far-infrared, as well as Raman-Balan states that “the Raman technique possesses special advantages, including being nondestructive; fast to acquire; capable of providing information at the molecular level and analyzing samples in aqueous solutions since water produces a weak Raman scattering; and important in the biochemical field where researchers study the ionization behavior, pH change, or amino acid configuration.” We chose Raman after carefully considering our target-blood in motion when approached transcutaneously, nanomolecules - safety of the laser when applied near the skin, and the already known Raman fingerprints of multiple infections and molecules in specimens, such as sputum and saliva.

Using Raman and machine learning (ML), the pathway from a blood-borne molecule(s) to a diagnostic prediction is as follows: A monochromatic laser (we used 830nm) is directed to the finger (not touching). An exchange of energy between the laser and the blood-borne molecules results in a scattering of photons in a way dependent on the particular molecular structure and bonds. The difference in wavelength of the scattered photons compared to the laser source is referred to as the Raman shift. After the inelastically scattered photons pass through a series of filters, a detector (back-illuminated CCD in our case) measures the light, which is converted into a spectrum. The Raman spectrum is a fingerprint that gives information about the molecules present in the sample (transcutaneous blood in our case) by analyzing the position, height, and width of peaks in the spectrum. Raman spectroscopy can identify lipids, proteins, nucleic acids, and amino acids within biological samples.

Machine learning (ML) has been combined with Raman spectroscopy to analyze multiple sorts of specimens. But it has not been applied in the manner in which we describe- a human finger’s vasculature when looked at transcutaneously and non-invasively. Moises Roberto Vallejo-Perez et al applied the classifier algorithms multilayer perceptron and linear discriminant analysis for “early detection of bacterial canker of tomato” (10). Lihao Zhang et al applied a support vector machine (SVM) analysis for the “classification of breast cancers’’ (11). Katherine Ember et al employed a multiple-instance learning (MIL) approach in their machine learning for “saliva-based detection of COVID-19 infection”, in which they achieved an area under the curve (AUC) of 0.8 and a sensitivity of 79% (males) and 84% (females), and a specificity of 75% (males) and 64% (females) (12). Davide Brinati et al performed a feasibility study (7)that analyzed actual blood tests, focusing on variables “age”, “wbc”, “crp”, “ast”, and “lymphocytes” from 102 COVID-19 negative and 177 COVID-19 positive patients, and applied different machine learning algorithms to achieve an accuracy of 82%-86%. Using a Random Forest classifier, their best results were accuracy 82%, sensitivity 92%, PPV 83%, AUC 84%, and specificity 65%.

We describe the feasibility of using a unique Raman patient interface that, together with machine learning, visualizes the blood non-invasively and transcutaneously to detect COVID-19. This is accomplished by simply inserting a finger into a patient-interface device that comprises a 830 Raman probe. The machine learning component of the viral detector allows the Raman component to become a more powerful predictive test of COVID-19 than if the Raman were employed without machine learning. Raman spectra have unique spectral characteristics for target molecules. Choosing which molecule to target in COVID-19 diagnostics and how to target it became our first challenge. We knew that with a high signal to noise ratio (SNR) Raman system, we would be able to obtain Raman spectra of the unique combinations and concentrations of molecules present in the blood of COVID-19 patients. We would examine transcutaneously the “soup” in the blood. We were confident that we could, in principle, obtain a high SNR transcutaneously because the Raman peaks of glucose had been measured transdermally in vivo with swine subjects. Kang et al (13) calculated the subtraction spectrum between tissue and pure glucose solution and determined that a high degree of similarity exists in the glucose peaks. We collected Raman spectra from a finger’s vasculature transcutaneously and non-invasively and compared that data with concurrent polymerase chain reaction (PCR) test results. We, therefore, had transcutaneous Raman spectra together with nasal swab PCR results from a group of COVID-19 positive and a group of COVID-19 negative people. The initial data (195 patients) were submitted to a Support Vector Machine (SVM) in order to arrive at a model that predicted COVID-19 with a specificity of 0.88 and a sensitivity of 0.75. However, as the study progressed with the database becoming more varied with different viral variants, vaccine effects, and clinical presentations, we employed a Gated Boosted Tree classification to improve the sensitivity of the model to 0.80, while maintaining the specificity to 0.75 with 476 patients. From a clinical perspective, we reasoned that false negatives (sensitivity) are more harmful than false positives (specificity). With 389 outpatients in our study, the possibility that an unknown medical condition common to all outpatient COVID-19 positive patients would account for the unique spectrum assigned to COVID-19 and thus confound our results is extremely unlikely.

## Materials and Methods

A Raman system with a laser (830nm) and a Back Illuminated-CCD is used to transcutaneously capture data that correspond to a given area in a finger’s blood vessel. The captured data was inputted into a machine learning classifier that had been previously trained to distinguish individuals who have been found to test positive for disease, such as Covid-19, from those who have been found to test negative for that disease. The classifier outputted an appropriate indicator, providing the user with an immediate indication of whether SARS-CoV-2 was present.

The viral detector assembly includes a 830 Raman system with a fiber probe housed in a unique patient interface into which an individual inserts a finger to receive the laser excitation (figures 1-5). The 830nm laser is selected as an excitation source to reduce fluorescence interference. The system also employs a back illuminated CCD to increase system sensitivity. The 830nm laser is transmitted through a 105 micro core fiber to the probe which is collimated and shined on the fiber. The Raman signal is captured and guided through a signal fiber to the Raman spectrograph. The laser powered at 250mW was used to capture Raman Spectra for (3) sequential 20-second intervals. Both the raw data (including fluorescence) and processed data (pure Raman) were stored.

The following images and schematics represent the viral detector prototype with its unique patient interface. The components include an 830nm laser (Innovative Photonics Solutions, Inc.), BI-CCD camera (Andor), Imaging Spectrograph SRaman 830 W2 Innovations, Inc., and Raman mprobe 830 (W2 Innovations, Inc.).

### Images and Schematics

U.S. Patent No. 11,452,454

U.S. Patent No. 11,304,605

### Viral Detector

Human interface

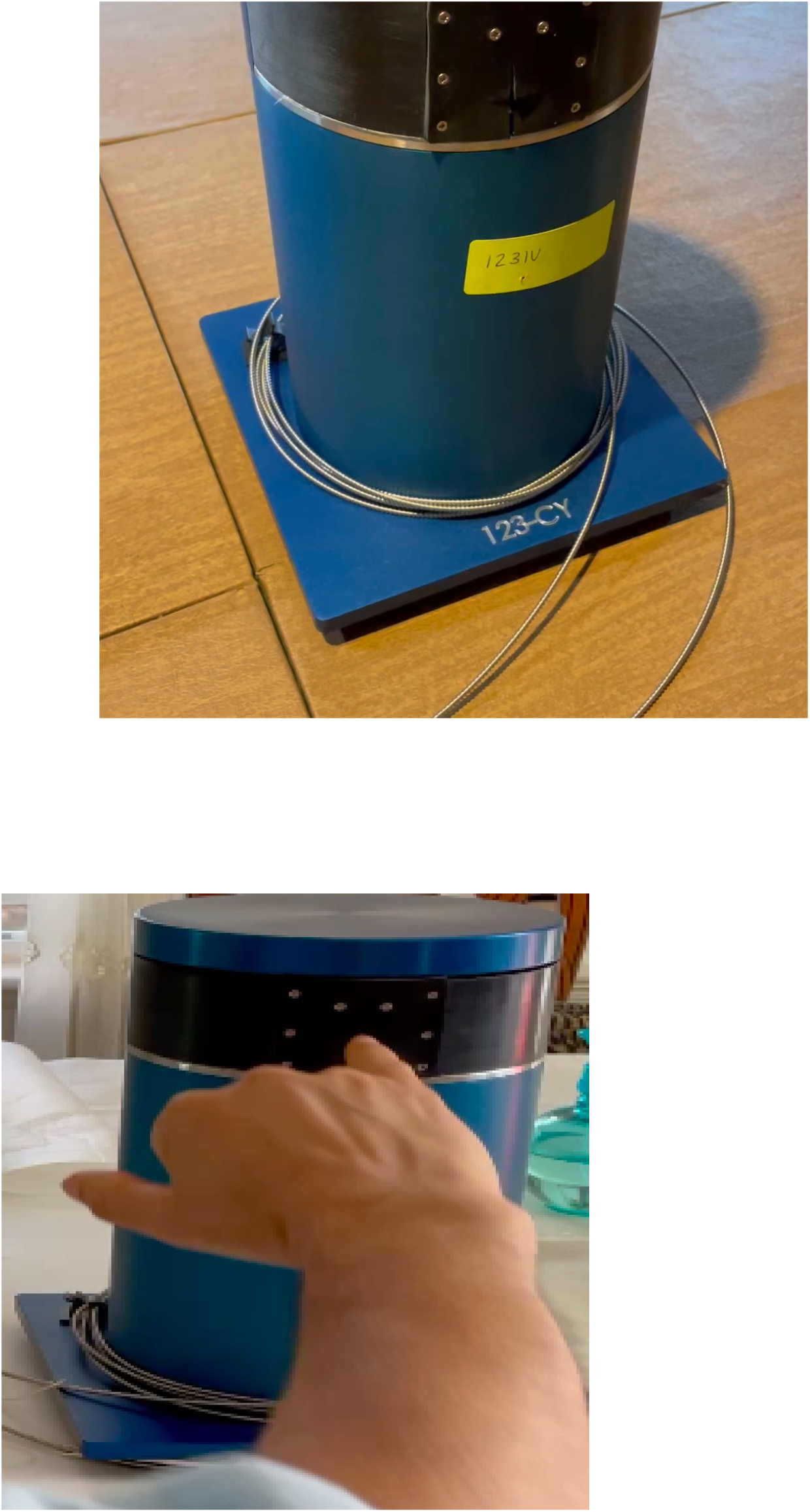

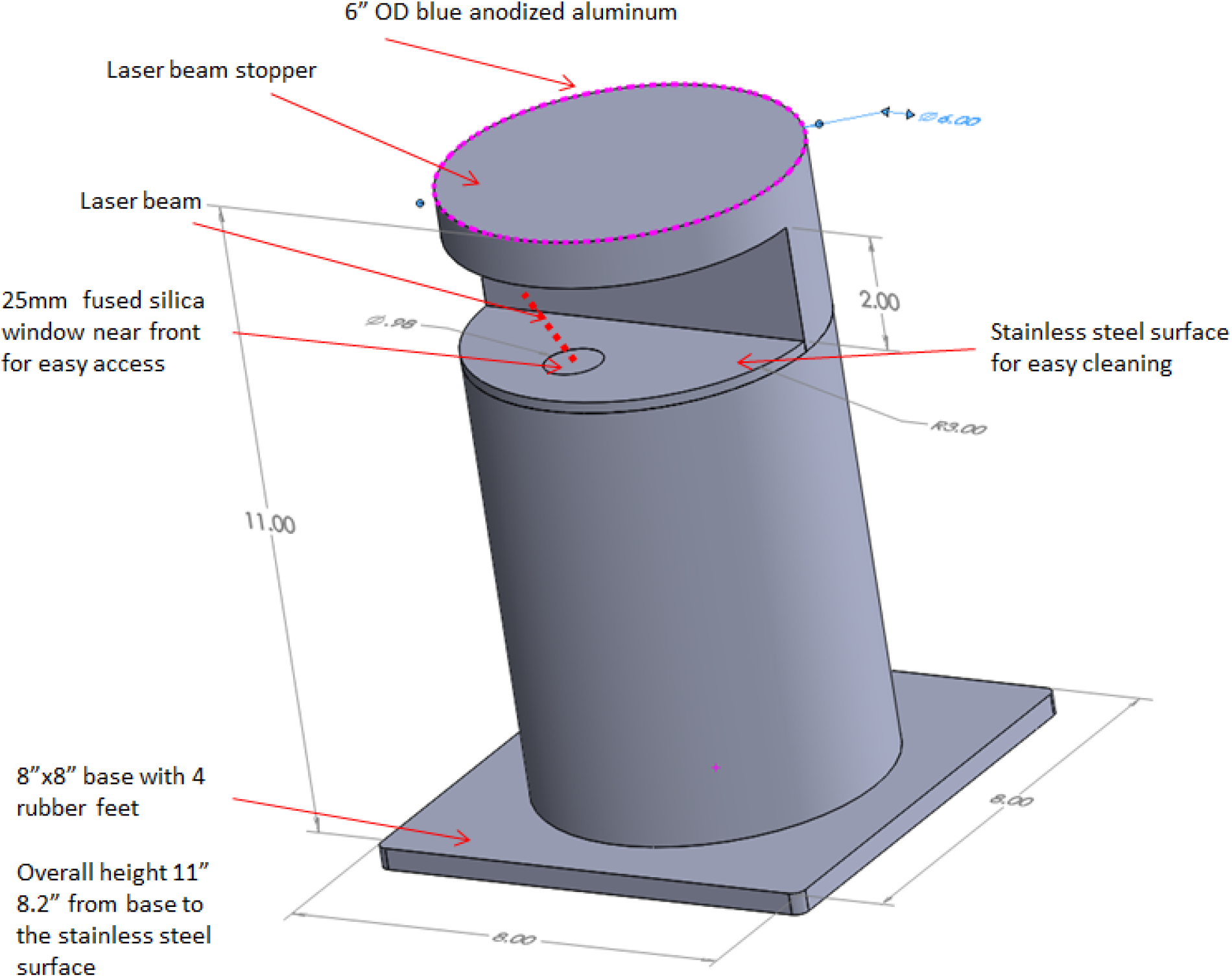

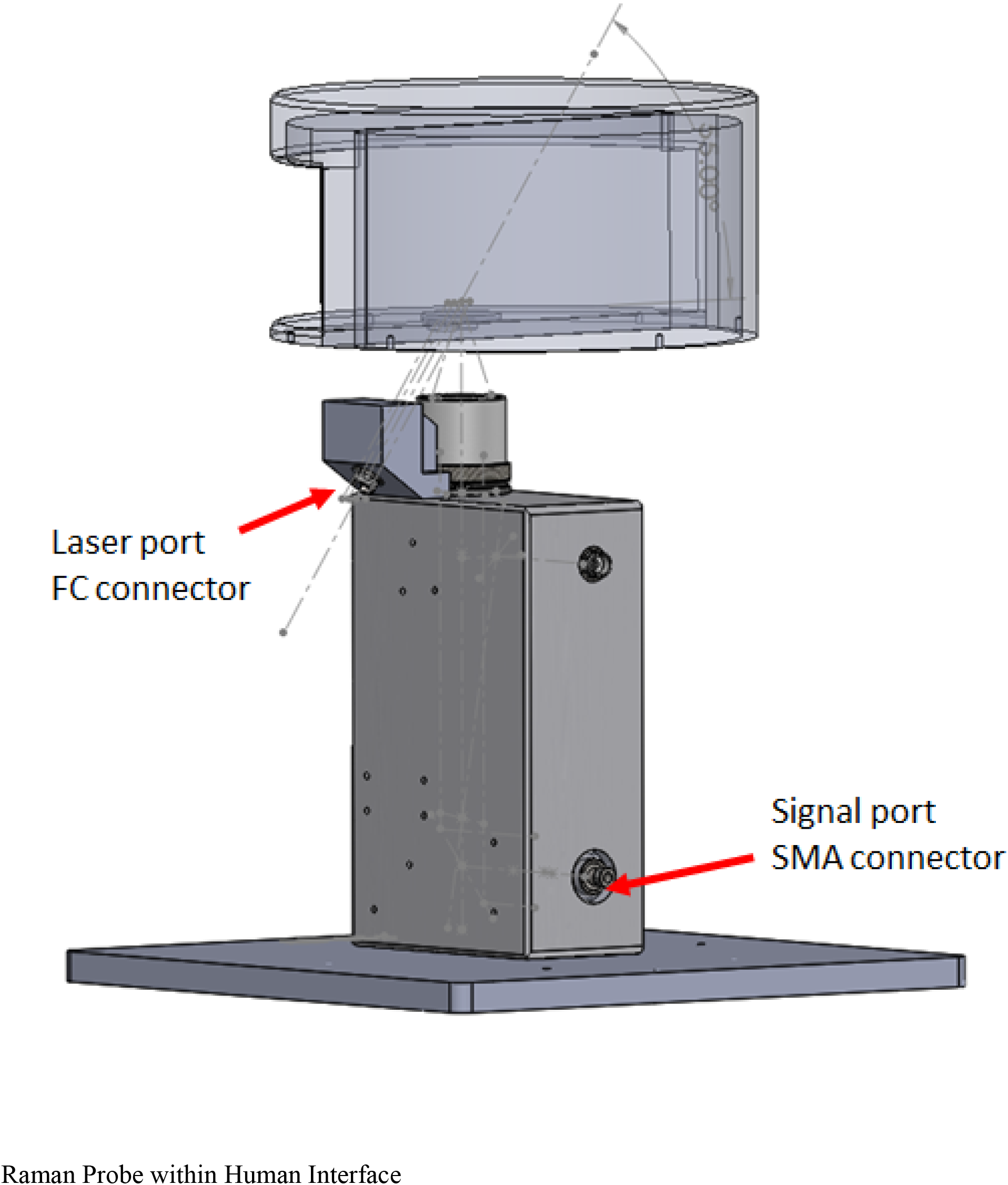

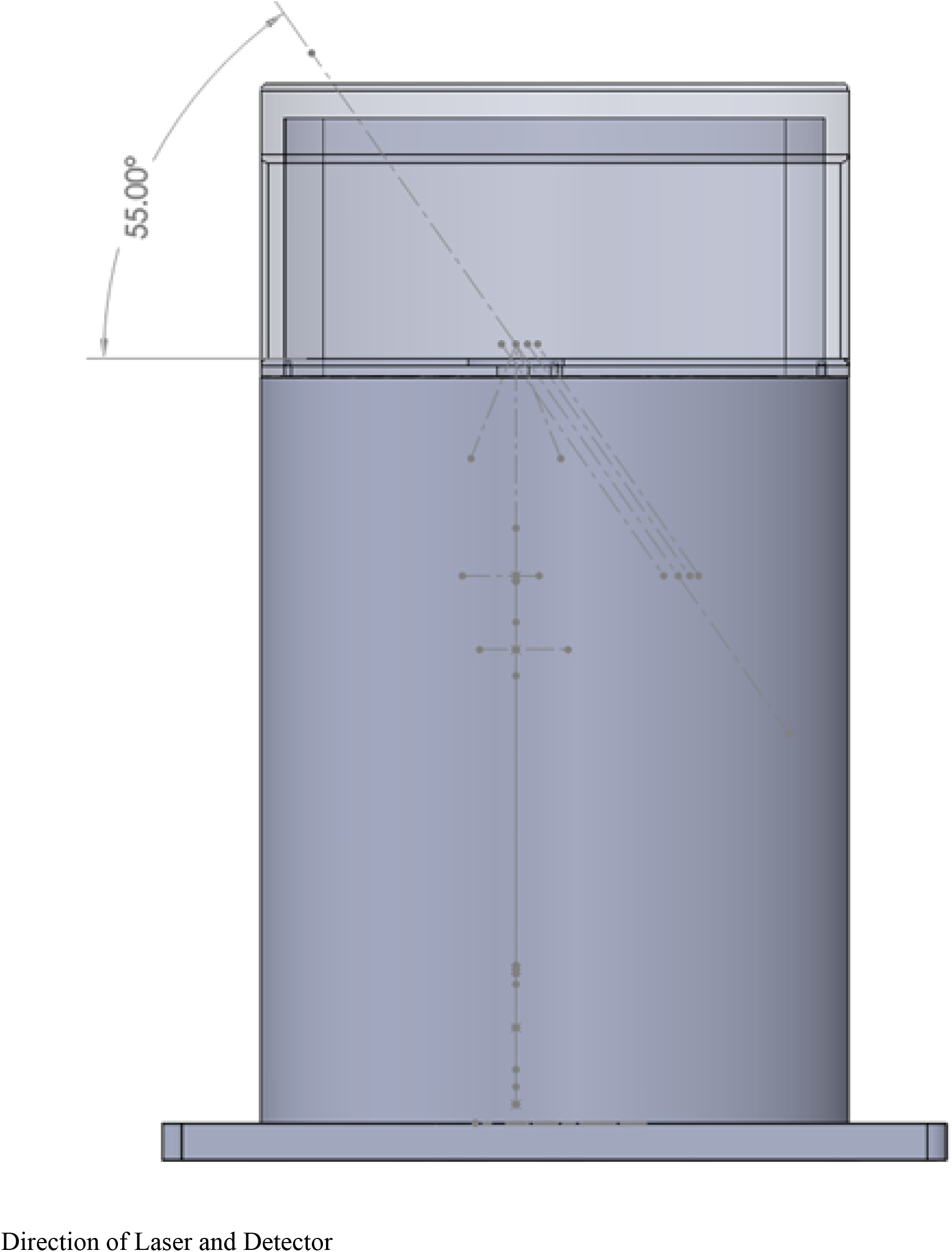

The device was inspected and the design analyzed for electrical, mechanical and laser safety. All electrical components are shrouded within a metal box and connections to the patient interface are fiber optic (not electric). A physical exam of electrical isolation was performed by using an ohm meter to check conductivity between the components of the system. The laser fiber sheath is conductive and is grounded. The patient interface is protected by the anodizing coating on the exterior. For use on patients, a medical grade isolation transformer was used to provide two means of patient protection per the requirements of 60601-1. The device uses a class 3B laser that is housed, including a permanent light blocking shield that conforms around the finger.

The viral detector device blocks a direct or reflected beam. Diffused reflections from a Class 3B laser such as those off paper or other matte surfaces are not harmful. The light blocking shield is made of black non-reflective surfaces and matte black paint was applied to the upper shiny surface of the device so that any reflections off the device itself are diffused. The modified device eliminates the risk of direct eye exposure and adequately mitigates any indirect exposure.

Direct measurements of the laser beam were taken at the patient interface without the light blocking insert in place. Measured laser power was below the Maximum Permissible Exposure (MPE) for Skin Exposure per ANSIz136.1 of 364mW/cm^2.

Prior to obtaining IRB approval at Holy Name Medical Center in Teaneck, NJ, the device underwent extensive safety testing at Sunrise Labs, Inc. in Bedford, NH. Informed consent was obtained for all patients. We excluded from the study anyone younger than 18 years of age. Patient data were collected for each patient, including age, symptoms, vaccination data, gender, and viral load (cycle numbers). Since transcutaneous devices have previously been documented to report incorrect results for non-caucasian skin colors, we also collected skin color data.

### Expected Outcome of the Research

We set out to demonstrate a correlation between positive and negative COVID-19 tests by PCR with the test results from the experimental transcutaneous COVID-19 detector.

### Exclusion criteria

Pregnant women and other special populations, such as minors and legally incompetent patients

### Recruitment

While undergoing PCR testing, either at the outpatient lab of Holy Name Medical Center or on the hospital ward, the finger test was done concurrently, as outlined in the protocol. No patient identifiers were entered. The PCR test was then processed at the Holy Name Medical Center lab, and results entered into the patient’s medical record. PCR test identifying numbers were recorded and stored.

### Study duration

The pilot study was conducted June, 2021 to August, 2022.

### Sample Size

In designing this feasibility study and its statistical power, we anticipated the need for 100 patients comprising a sample of 50 men/women with SARS-CoV-2 by PCR and 50 with negative PCR tests, according to the McNemar χ^2^ test. We actually studied 476 (316- and 160+) patients that we included in our data analysis.

### Variables

1. Raman Spectra
2. Inpatient/Outpatient
3. Symptoms
4. Fever
5. Cycle Threshold numbers
6. PCR results

### Secondary Outcomes

The collected data for each patient will allow subsequent studies and research to focus on outcomes not directly investigated with the current study. These include:

- effect of skin color on Raman measurements using our device.
- ability of our device to detect immune status
- ability of our device to calculate viral load
- Determining the presence of viral variants and predict nature of the variants.

Excluding factors such as age and gender, many of the above variables have blood-borne targets for Raman spectroscopy – antibodies, lymphocytes, and other inflammatory markers specific to COVID-19 – which provide unique Raman spectra.

### Ethics

This study protocol complied with the institutional review board (IRB) and Investigative Committee on Clinical Research (ICCR) at Holy Name medical Center and was conducted according to rules and guidelines of good clinical practice (GCP). Data was handled confidentially, and all data was stored for the length of the study and for 15 years afterwards at the site, for further publication.

Informed consent, approved by the Center’s Ethical Committee, was obtained from the subject by an authorized research team member only.

## Results

Data analysis confirms that the non-invasive viral detector(123-CY) has predictive power and is able to distinguish COVID-19 positive from COVID-19 negative patients. For the initial 195 patients, on visual inspection alone of the Raman spectra, a distinct difference was noted from 860-1100 cm^-1^ Raman shift (see graphs 1,2,3). For COVID-19 positive patients, there were 3 peaks, all of similar amplitude. For COVID negative patients, the smaller middle peak is flanked by 2 higher peaks. However, we recognized that visual inspections alone of graphs can be deceptive. When the text files were subjected to computer analysis, 1000 data points per file, the differences between COVID-19 positive and negative files were significant. As more data was inputted and analyzed with “Gradient Boosted Trees” (GBT) classification, the predictive power increased. Including both asymptomatic and symptomatic positives (N=70) with negatives (N=125), the sensitivity was 0.75 and the specificity was 0.88 for that initial group of 195 patients. The positive predictive value was 0.74, and the area under receiver operating characteristic (AUROC) was 0.81 (see graph 4). However, with increasing data input and algorithmic analysis, as opposed to simple visual inspection, we detected additional significant differences in the Raman Spectra of positives and negatives in the 1700-2200 cm-1 range. What this study strongly suggests is that with increasing data, the prototype- a non-invasive, immediate, trans-cutaneous viral detector-becomes more powerful.

The following Raman spectra obtained from our device, whose text files were processed with machine-learning, are representative of the high SNR of our working prototype.

### Raman Spectra: Transcutaneous, Non-invasive (Finger)

Same patient tested using different fingers:

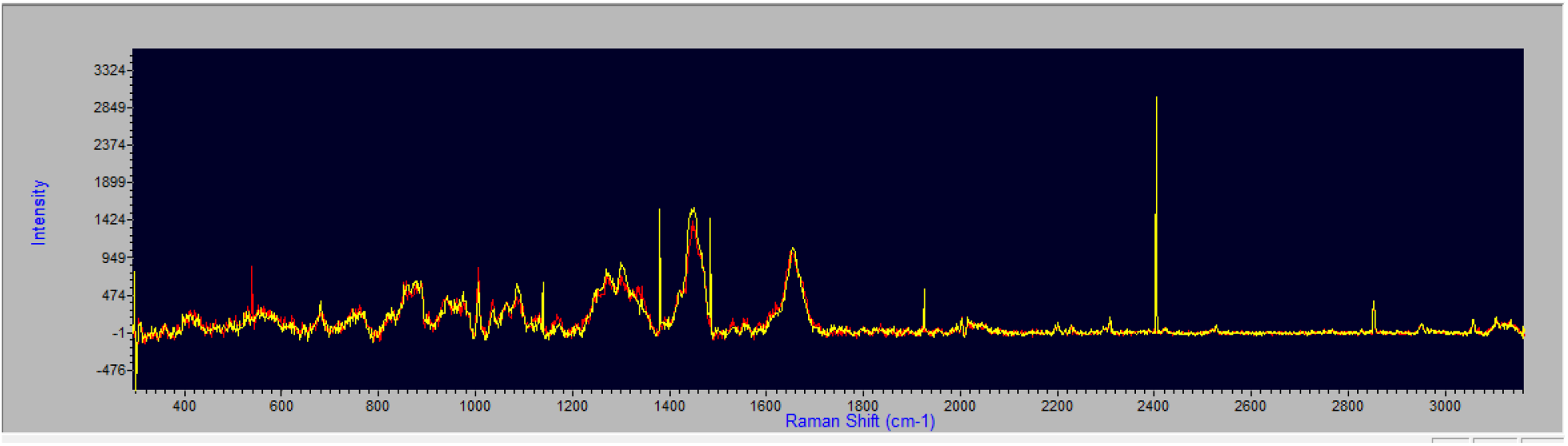

COVID-19 Negative Spectrum: Processed

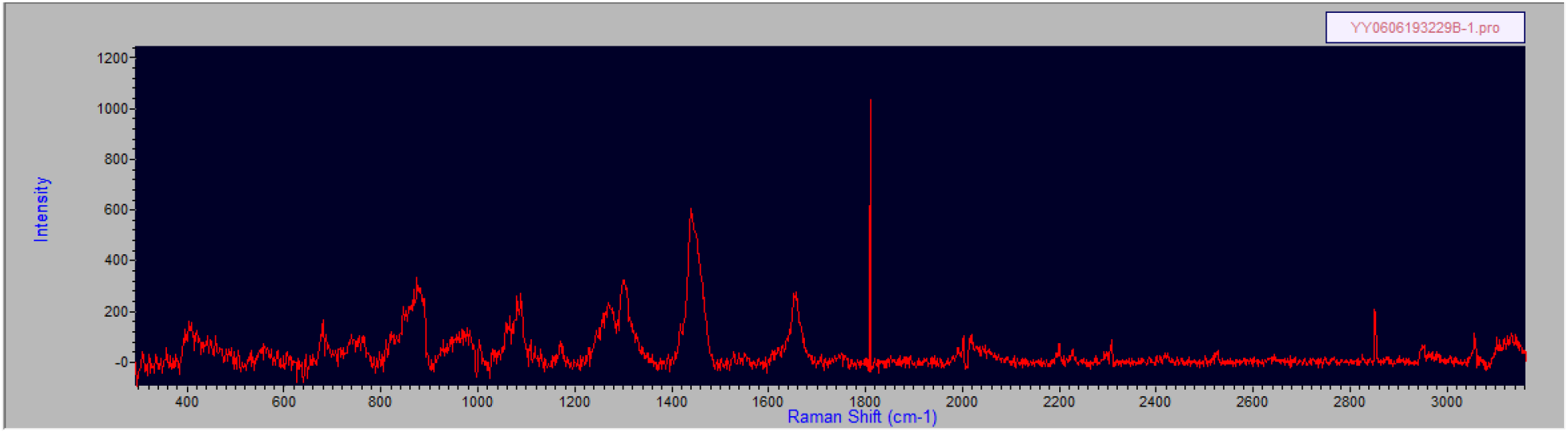

COVID-19 Negative Spectrum: Raw

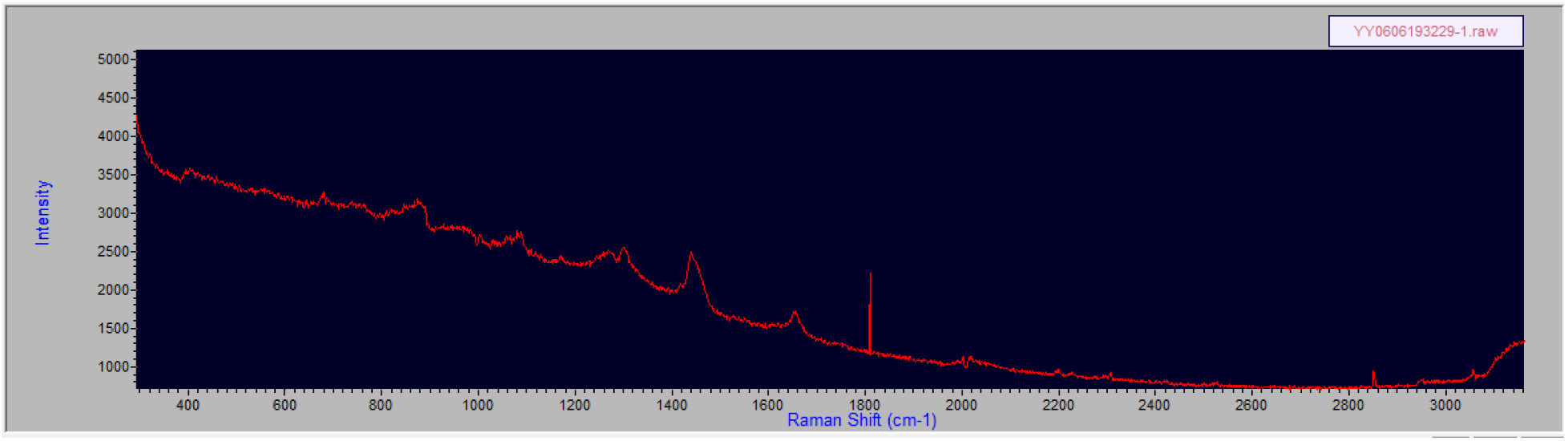

COVID-19 Negative Spectrum: Processed

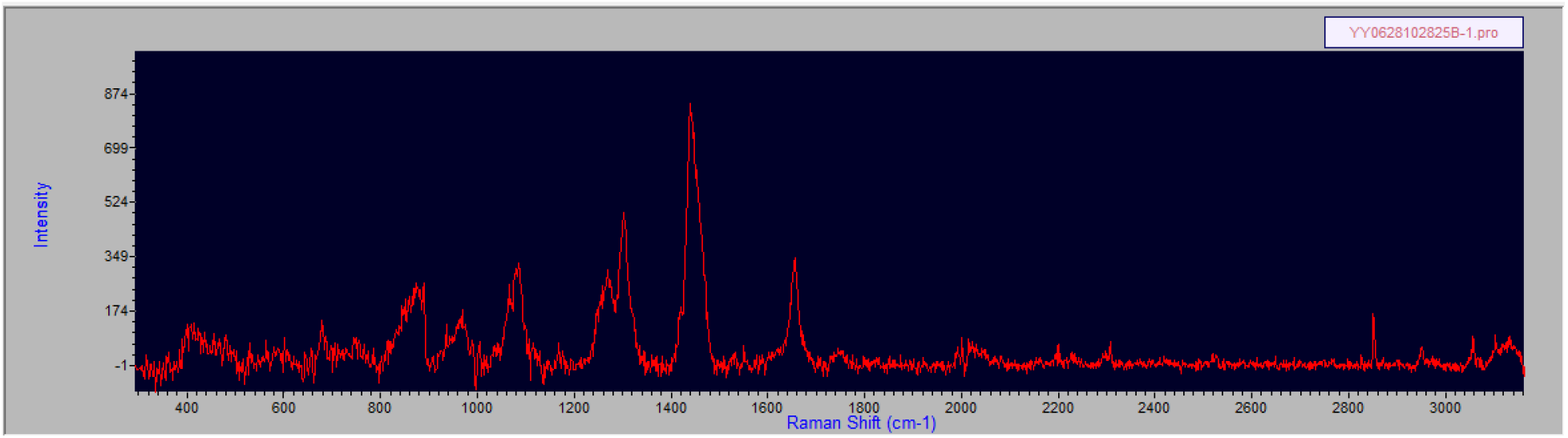

COVID-19 Negative Spectrum: Raw

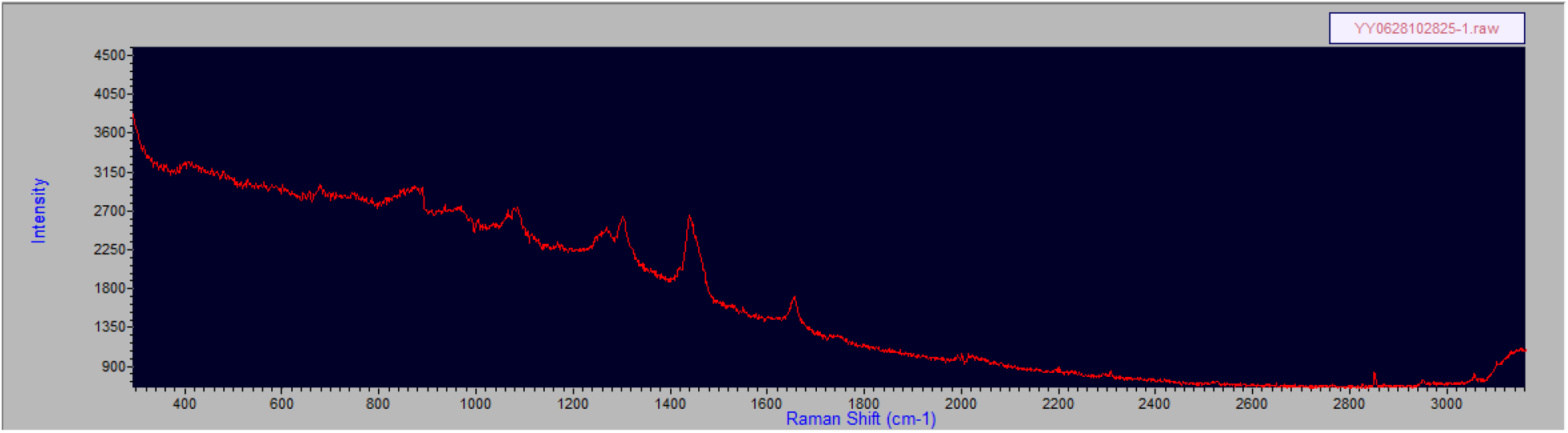

COVID-19 Positive Spectrum: Processed

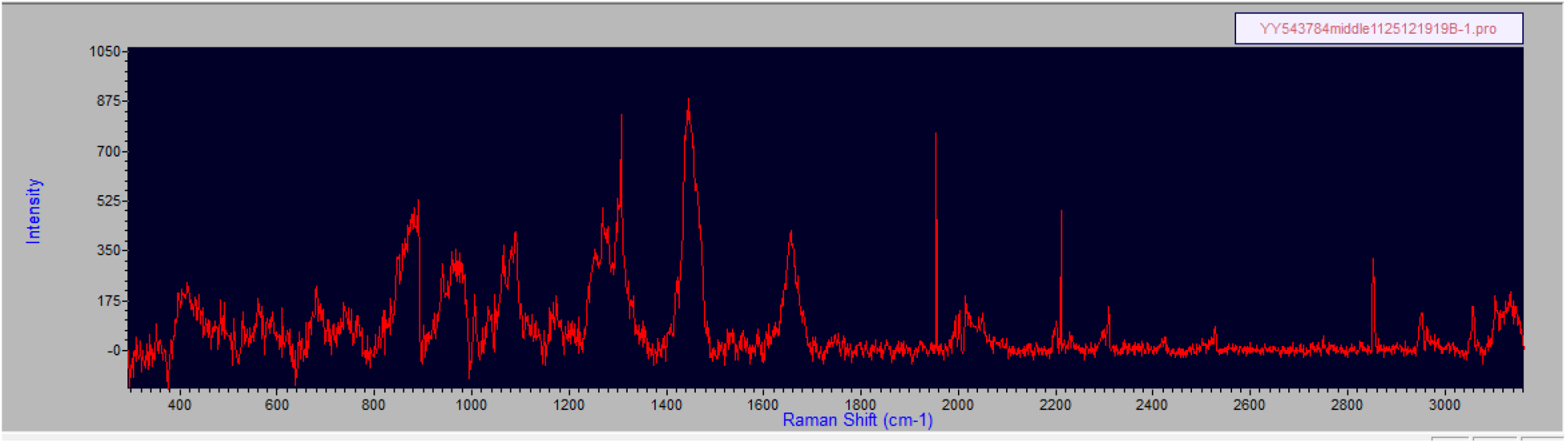

COVID-19 Positive Spectrum: Raw

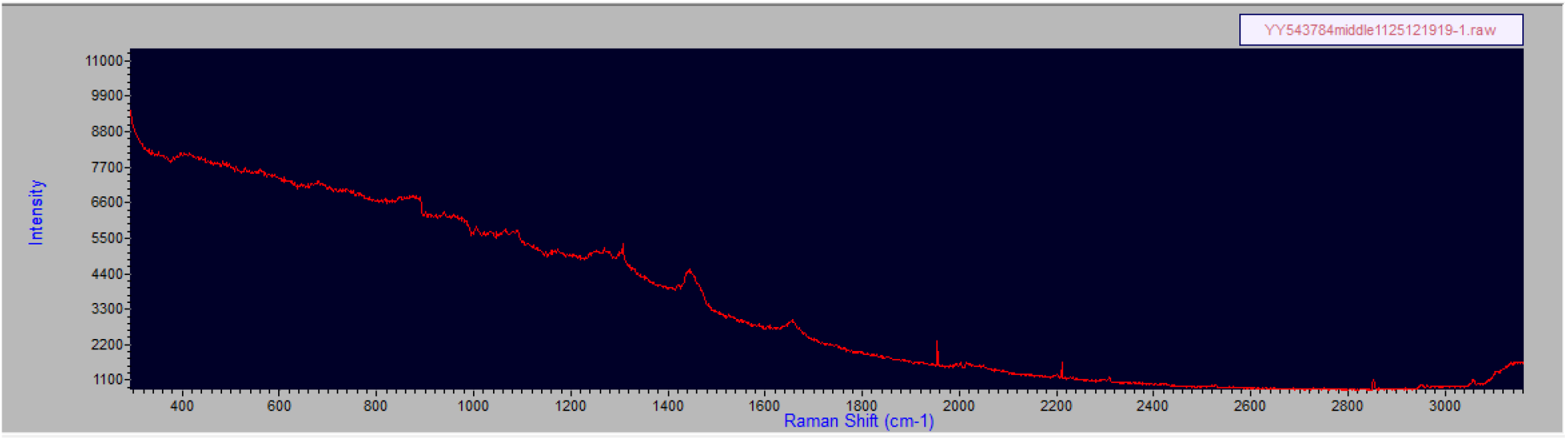

COVID-19 Positive Spectrum: Processed

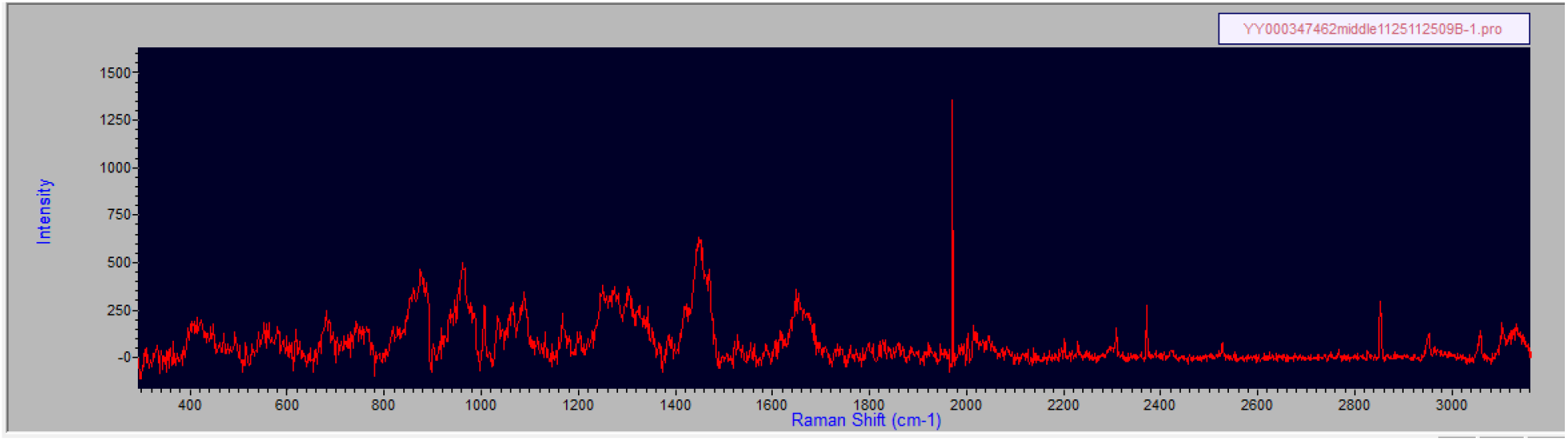

COVID-19 Positive Spectrum: Raw

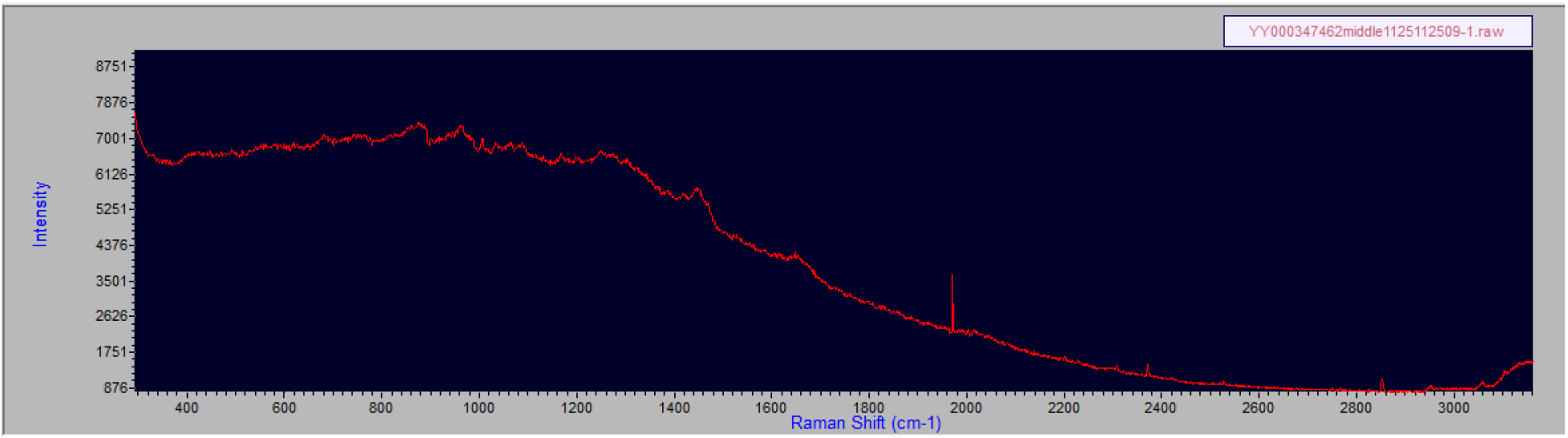

COVID-19 Negative Spectra from 4 patients:

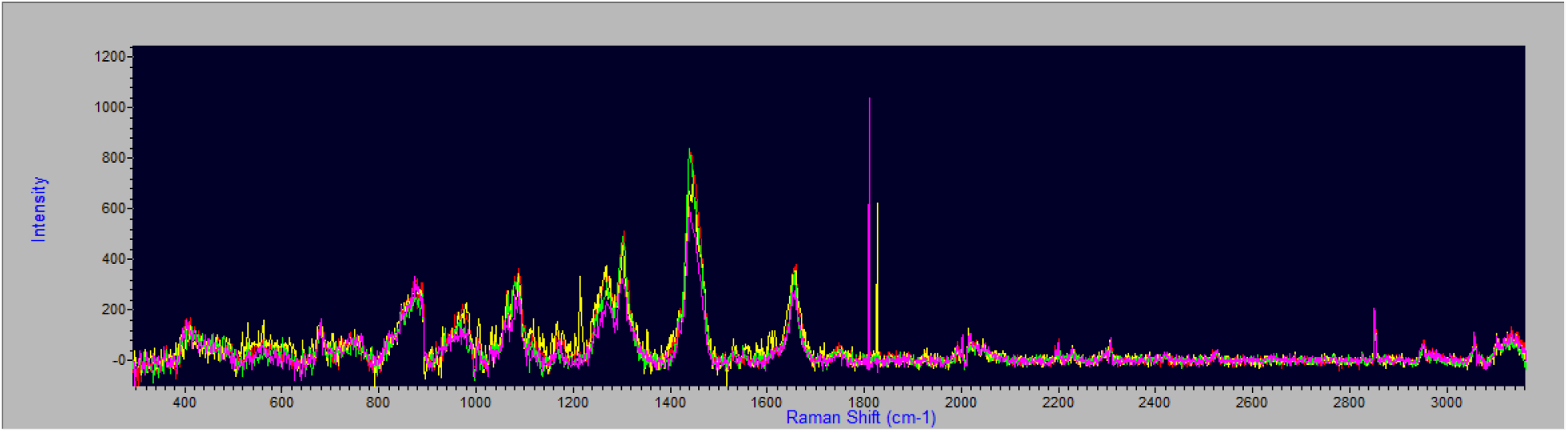

COVID-19 Positive Spectra from 3 patients:

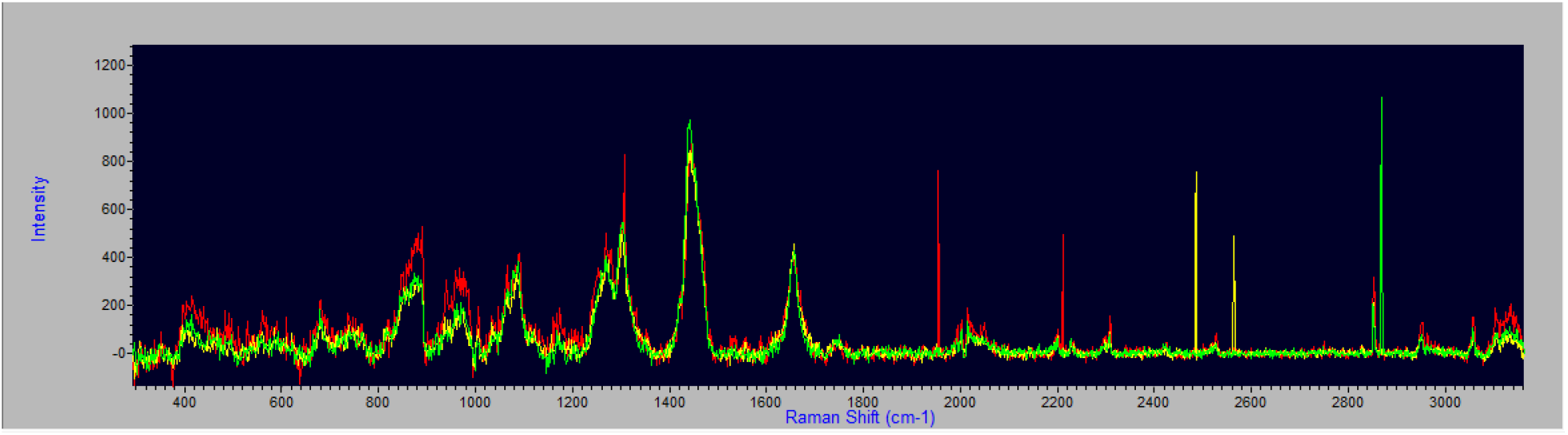

COVID-19 Spectra Positive and Negative Overlap: Pos(yellow and red) and Neg(green and purple)

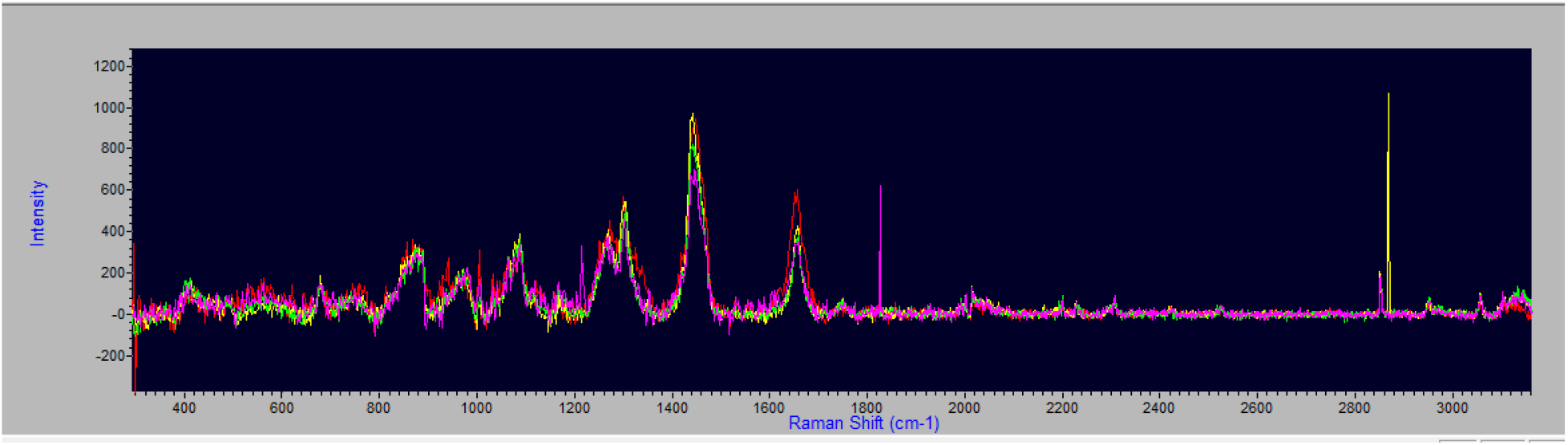

From Carlomagno et al:

### Raman Spectra: Extra-Corporeal Specimen

**Positives:** 860-1100—more gradual upslope first peak, and middle peak closer in amplitude to 2 flanking peaks. 2 superimposed spectra from 2 COVID-19 positive patients

**Negatives**: 860-1100- sharper upslope and greater difference in amplitude between middle peak and 2 flanking peaks in COVID-19 negative.

Graph 1:

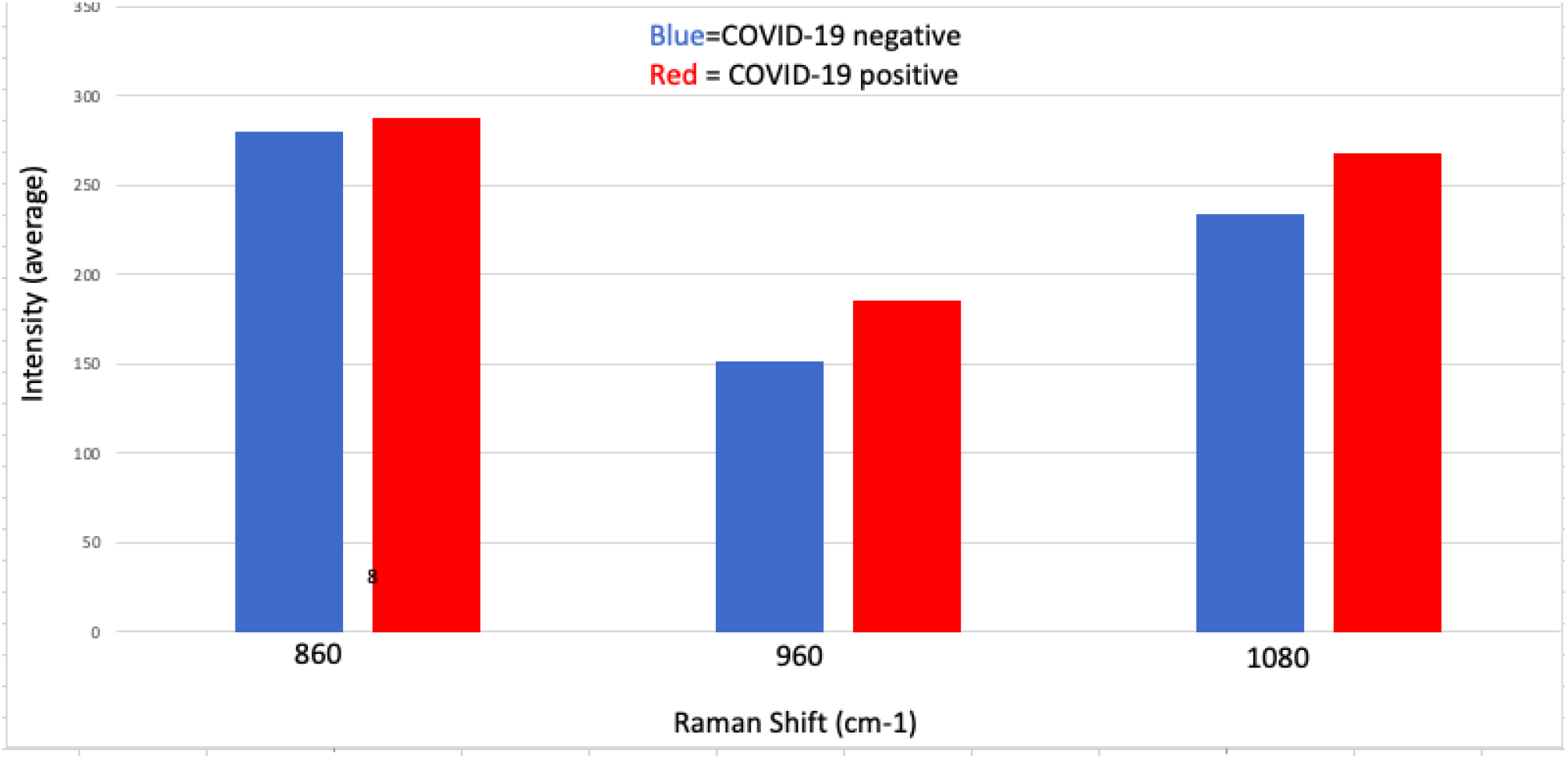

Graph 2:

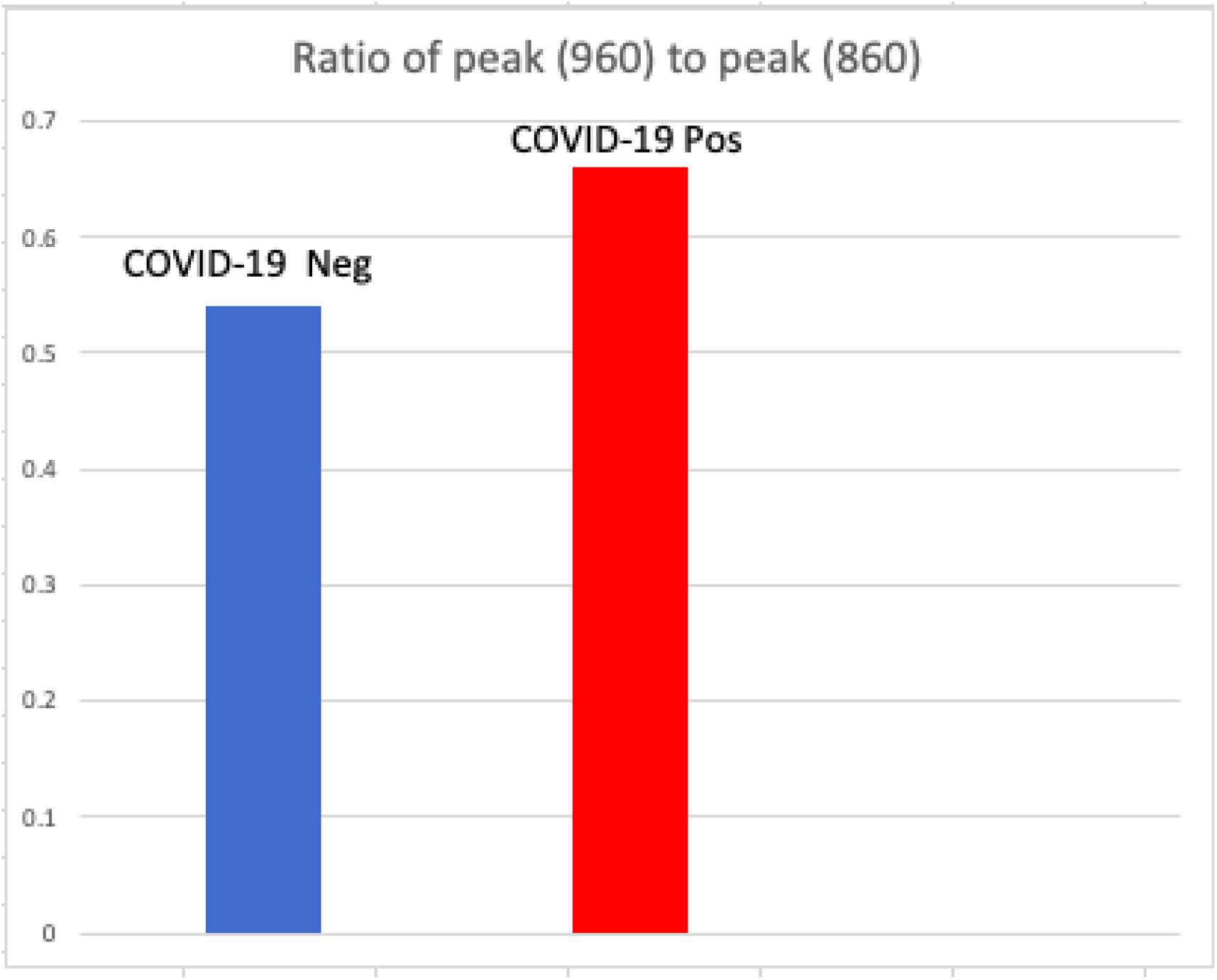

Graph 3:

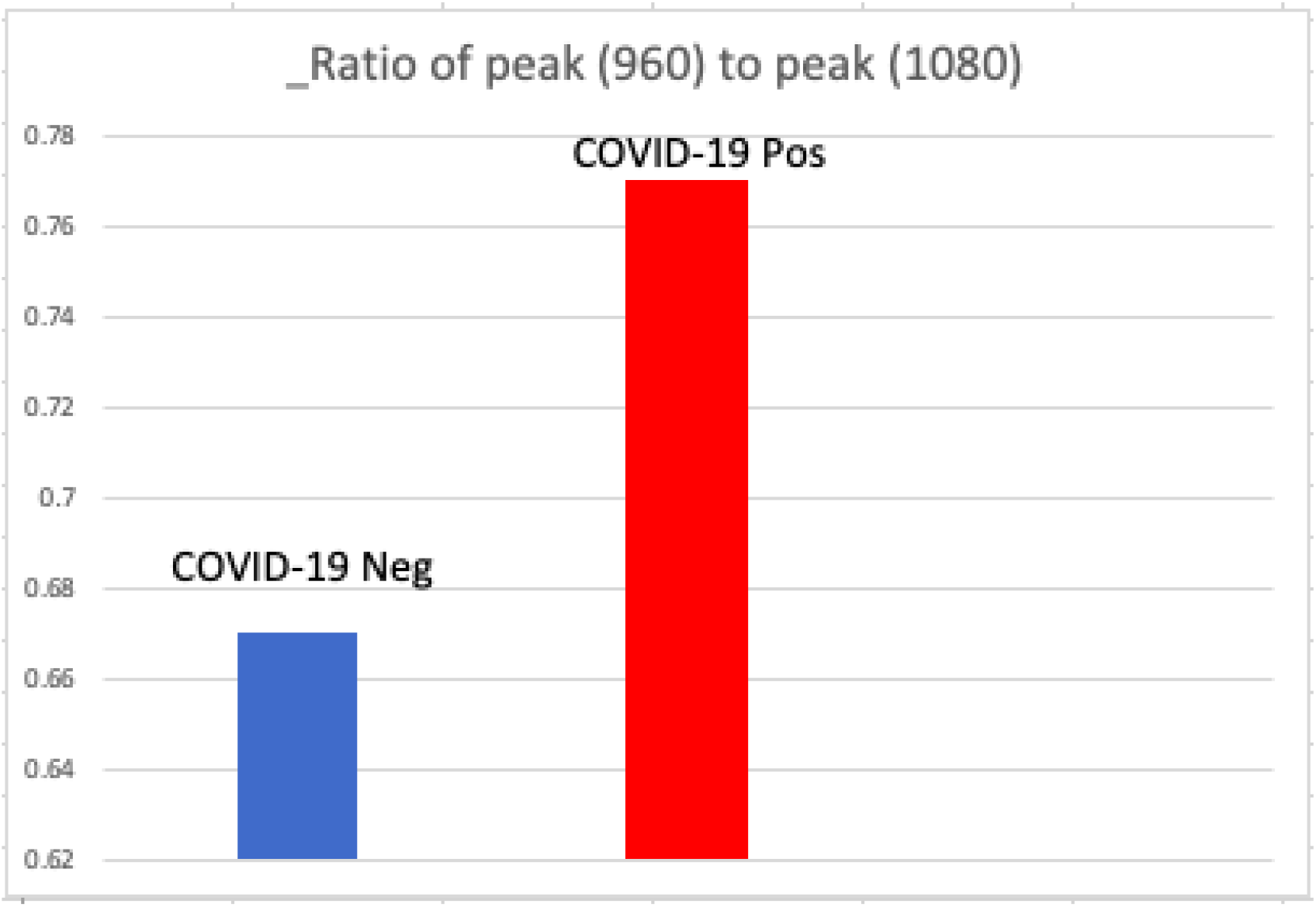

Graph 4:

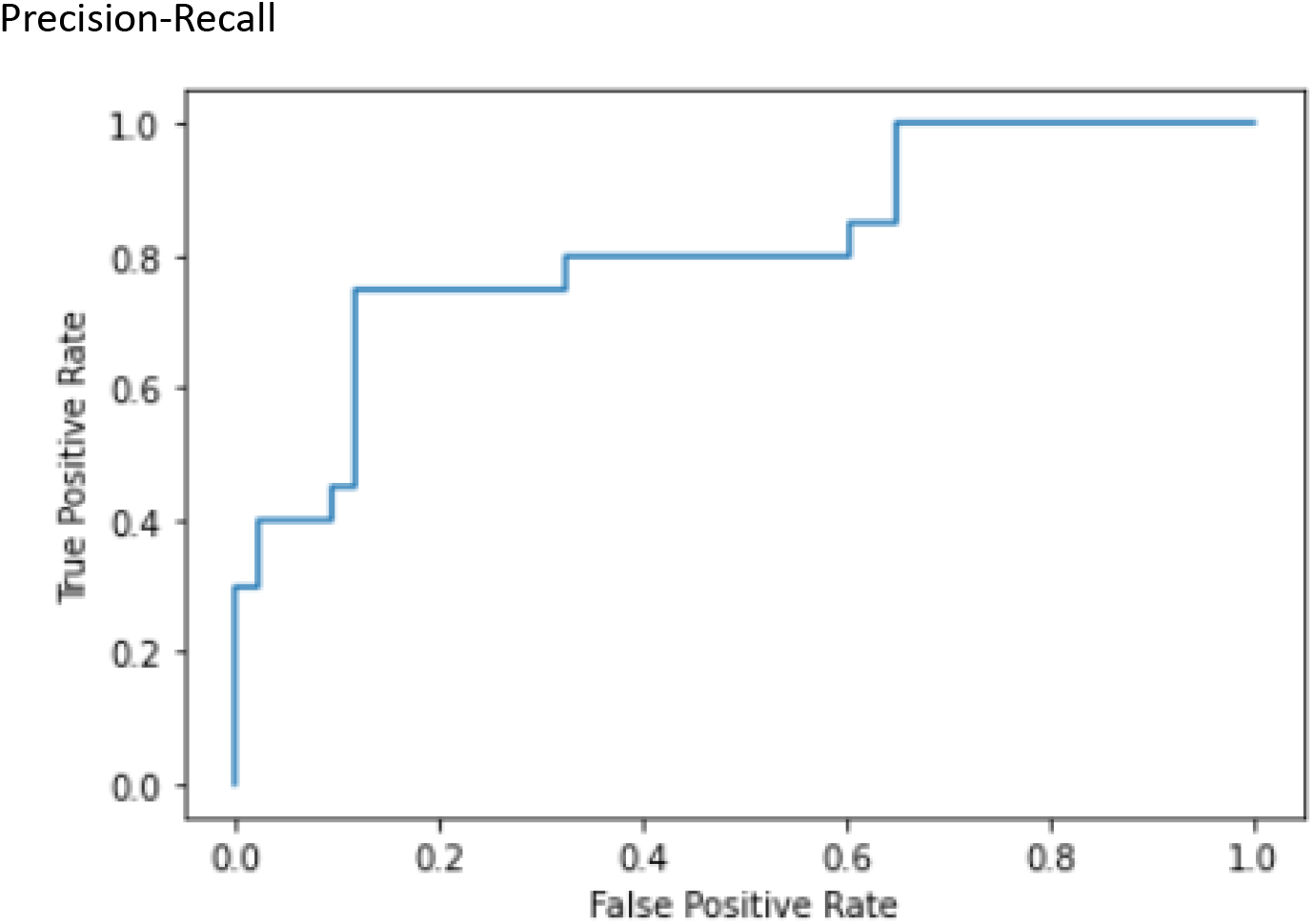

**Fig.1:**
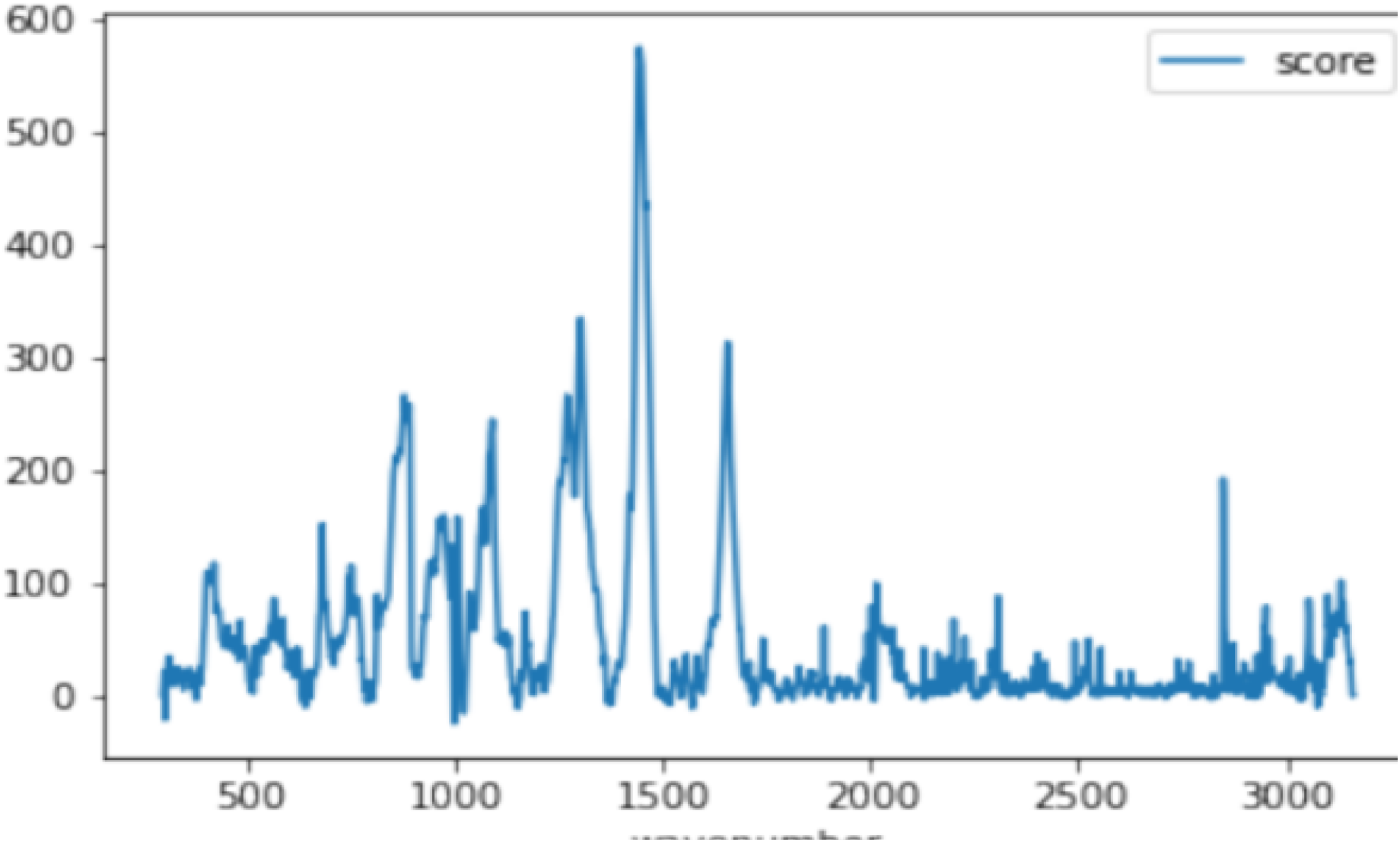
Average Raman spectrum output from the device (average intensity shown across cases and controls in the training dataset)

## DATA ANALYSIS

Using a python engine, we matched text files data with PCR results. Two random splits of the data were created as (train) and (test). A preliminary analysis using t-tests per wavenumber of Raman spectra was obtained- see Fig. 2 above. This suggested that individual peaks contained information to help distinguish cases and controls. Fitting a machine learning model with data setup then ensued. The data was set up as a pivot. The Raman score was treated as a feature so each patient corresponded to a 2000-dim feature vector. In the training set, we fitted a StandardScaler to standardize each feature. The same scaling was applied to the test set. A gradient boosted trees classification to train and model to test on keepaway data was performed. A Precision-Recall Curve was generated, as shown above.

**Fig.2:**
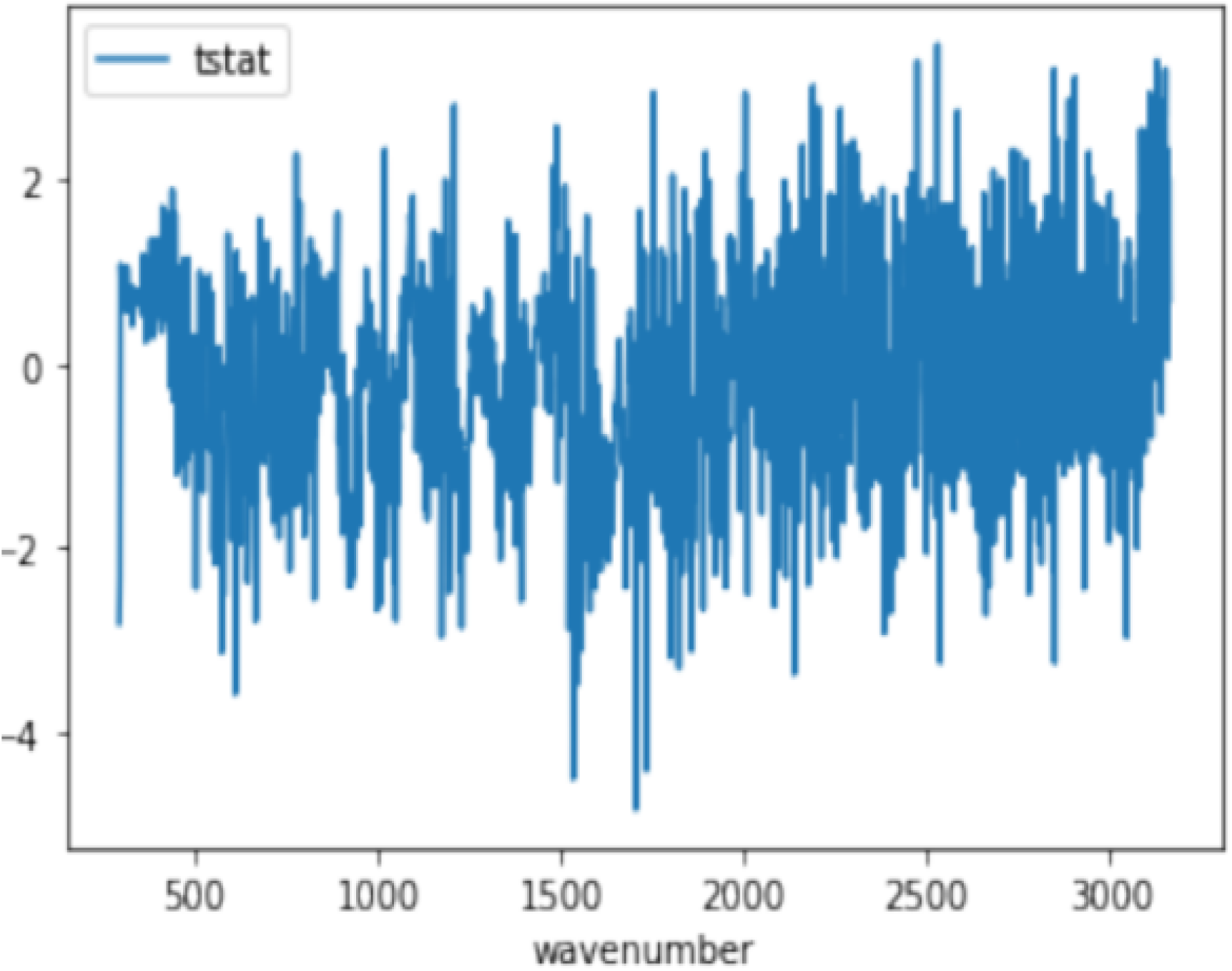
Individual Raman peaks contain information that can distinguish between cases and controls T-tests on score z: For each individual, wavenumber values (across the 2000 points) were converted to z-scores to control for patient-specific biases.

### Machine-Learning

The following provides details regarding supervised machine learning, which is the technique used in the learning approach herein. Supervised learning is the machine learning task of inferring a function from labeled training data. The training data consist of a set of training examples. In supervised learning, typically each example is a pair consisting of an input object (typically a vector), and a desired output value (also called the supervisory signal). A supervised learning algorithm analyzes the training data and produces an inferred function, which can be used for mapping new examples. An optimal scenario allows for the algorithm to correctly determine the class labels for unseen instances. This requires the learning algorithm to generalize reasonably from the training data to unseen situations.

The following steps were used. An initial determination of what kind of data was to be used as a training set (total 125 negative and 70 positive examples as of 1/15/2022, with a train-vs.-test split of 67-33%). Here, the training data included Raman spectra captured from patients using the device configuration described above. The training set was then gathered. In particular, during the training phase, the absorption spectrum was measured, quantized, and digitized (intensity measurements for 2000 wavenumbers). We constructed a vector for each patient.

The corresponding outputs were gathered from measurements using the described device (each observation corresponded to an individual and the corresponding output represents a binary variable corresponding to the status of the individual’s Covid RT-PCR test). Then, an input feature representation of the learned function was determined (transforming each of the 2000 features to a standard form: a mean of zero and variance of one). In this approach, the input object was transformed into a feature vector, which contains a number of features that are descriptive of the object. The structure of the learned function and corresponding learning algorithm were then determined. The learning algorithm was then run on the gathered training set. Some supervised learning algorithms require a user to determine certain control parameters (for the gradient boosted machines, we set the number of tree-classifiers as 500). These parameters may be adjusted by optimizing performance on a subset (called a validation set) of the training set, or via cross-validation. The accuracy of the learned function was then evaluated. After parameter adjustment and learning, the performance of the resulting function was measured on a test set that was separate from the training set (sensitivity/specificity 75%/88% for gradient boosted machines).

The power of this diagnostic device with its integrated AI and the increasing accuracy with increasing data input were demonstrated when the data analysis was performed on additional patients numbering 476 patients total. The sensitivity of the test increased to 0.80, while the specificity remained above 0.75.

## Discussion

We describe a viral detector that employs Raman spectroscopy with a unique patient interface and, using specific hardware parameters together with machine learning, can predict COVID-19 transcutaneously and non-invasively. We were able to prove that when our device targets the blood transcutaneously, the derived Raman spectra together with machine learning produces a significantly different result between COVID-19(+) and COVID-19(-) patients. We included in our study both inpatients and outpatients, symptomatic and asymptomatic.

In conceiving our idea and unique design for the viral detector, we anticipated various challenges that were sufficiently met. Although COVID-19 is a “respiratory” virus, we believed at the outset that COVID-19 could be detected by investigating the blood. We reasoned that the rich vasculature of the aero-digestive tract, upper and lower airways, and oropharynx would absorb almost anything inhaled, as occurs with many inhaled substances. Even if the virus itself were not in the blood, or at a concentration too low to reliably detect with our device, the increasing reports (7,8,9) of systemic effects of COVID-19 were strong evidence that there were hematological abnormalities to target (9). We also assumed that with adequate laser power, set at the proper wavelength, at a proper distance and angle from the inserted finger using no blood draws, we would be able to detect the Raman signature of COVID-19-- the precise spectrum of the unique combination of parameters in the blood that distinguishes COVID-19 positive from negative patients.

We chose the specific input variables-PCR results, Cycle Threshold, Inpatient vs. Outpatient, Fever, Symptoms- as these variables are routinely obtained in a rapid “drive-through” testing center. We could have added other variables, such as Oxygen Saturation, which may have increased our test’s accuracy further. However, in the interest of speed and convenience, we did not.

We did not assume that an enclosed laser in our viral detector device would necessarily be safe for human use. Thus, we first designed the device and adjusted the parameters to minimize any potential risk ranging from finger burns to eye injury. We then enlisted the expertise of an independent third-party laser safety testing company to run a battery of tests that confirmed the device’s safety.

Our pilot study enrolled 476 patients, none of whom experienced any adverse effects from the device. We anticipate that we can improve our results with further modifications of the device’s parameters, as well as more data input.

Raman spectroscopy is named after C.V. Raman, a physicist who observed the inelastic scattering of light in organic liquids in 1928. The instruments typically include a light source, such as a laser, a spectrograph to collect and disperse the scattered light by Fourier Transform methods, a detector, such as a CCD, and a filter to separate the Raman scattering from other light signals.

Raman spectroscopy detects precise vibrational modes of molecules and, as such, provides a structural fingerprint of molecules. Its widest application, especially until 20 years ago, was in the chemical analysis of molecules, such as counterfeit drugs, pharmaceuticals, and many other applications. With the advent of Spatially Offset Raman Spectroscopy (SORS) (3,15) and other deep tissue techniques, intense interest and ground-breaking work ensued. More attention was being aimed at biomedical applications (3,4). However, those applications of Raman typically included a specimen on a slide, other platform, or container (16), or a direct application of a probe to an extra-corporeal specimen (16,17,18). Our objective was to safely combine the best Raman parameters of laser power, wavelength, distance, detector, and spectrograph processing with the most powerful machine learning algorithm in order to non-invasively “see” beneath the skin into the human vasculature and target the many molecules that represent the intravascular unique signature combinations, both quantitatively and qualitatively, in an infected Covid-19 patient.

The underlying biochemistry of the expected acute phase reactants, as well as viral composition, has been studied using Raman Spectroscopy. Whether using ex-vivo blood or purified lab samples of various acute phase reactants, the Raman Spectra for many of these molecules has already been discovered and has served as the scientific basis of our study.

For IL-10, Shuo Zhang et al (22) found peaks at 852, 1064, 1298 and 1470 cm^− 1.^

“The peak at 852 cm^− 1^ might be from proline, hydroxyproline and tyrosine, 1298 cm^− 1^ is suspected from CH_2_ deformation modes and 1470 cm^− 1^ can be assigned to a C=N stretching bond within the IL-10 protein structure. For the spectra of IL-10, the SNR of highest peak at 1064 cm^− 1^ is 5.02. Only a single peak at 990 cm^− 1^ is found in IL-21 spectra with SNR of 6.10.”

O-Cuevas et al (23) describe the Raman spectrum of IL-6 showing bands at 938, 1261 and 1655 cm-1 due to the C–C stretching of the backbone structure, amide III and amide I, respectively, corresponding to the typical a-helix secondary structure conformation of IL-6. In addition, conventional residues of amino acids, such as the band at 708 cm-1, indicates the C–S stretching of cysteine residues in the IL-6 structure. Therefore, the conformational structure of IL-6 was corroborated using Raman spectroscopy.

N Chaudhary et al (24) describe the Raman spectra for monocyte and lymphocyte subgroups, as well as IL-4.

For C-Reactive protein, Bergholt (25) collected plasma from 40 individuals and with high sensitivity identified the molecular structure of C-Reactive protein using 785nm excitation and Raman spectra.

Staritzbichler et al(26) analyzed 234 blood samples for 38 biomarkers using Raman spectroscopy at 785nm excitation. They included creatinine and cystatin C as markers of renal function and C-reactive protein (CRP) and interleukin 6 as biomarkers of inflammation.

They also included the patients’ results for the following biomarkers in their analysis, although these were measured as whole blood samples or hemolysate and are not detectable in serum: glycated hemoglobin (hemoglobin A1c, HbA1c), blood cells (platelets, white blood cells), and international normalized ratio (INR)).

In our study, we examined, non-invasively, the entire mixture of blood components that when viewed as a whole can predict the presence or absence of COVID-19. The naked eye suggested a subtle difference, from 860 to 1100 cm-1 Raman shift, between positives and controls. The text files from the Raman spectra, when processed with machine-learning, detected differences between COVID-19 positives and negatives. At 2200, 2400, and 1800 cm-1, differences were detected. Glycerol, a metabolic product of glycerides and lipids, is known to produce a peak in this area (1000-1500) when examined on a slide. It is possible that a change in lipid metabolism occurs as a result of COVID-19, similar to the alteration in the clotting cascade. Our differences in Raman spectra between positives and negatives would be consistent with the findings of Roberts et al (19) in their study of the metabolomics of COVID-19.

Activity at **1700** Raman shift is strongly associated with molecules containing a carbonyl group **C=O**, such as proteins. In fact, carbonyl groups are found in HIV as well as COVID (20). Higher shifts in the **2000** range are associated with cyano groups **C=N**, also in molecules such as RNA. Perhaps we are detecting viral breakdown products (spike protein and RNA), as well as some other acute phase proteins.

Further study is necessary to assign a particular Raman peak to a specific molecule in our spectrum detected transcutaneously. Because we combined machine learning with spectroscopy, and since the anticipated concentration of any one particular molecule unique to COVID-19 is low, it is precisely the combinations of molecules we sought to detect and analyze. Applying Raman transcutaneously for diagnostic purposes has not been established. We may infer from saliva or other specimens placed on a slide which particular Raman peak corresponds to which molecule associated with COVID-19, such as the many acute phase reactants and viral RNA, when applied transcutaneously. However, our aim with this proof-of-concept study was to analyze the complete picture or Raman spectrum using machine learning algorithms.

In designing the prospective, observational study, as well as the basic science behind the device, we anticipated any potential bias. We compared our diagnostic device with the gold standard RT-PCR, performed in a hospital approved lab. As we classified in our data 87 inpatients as well as 215 outpatients who tested positive for COVID-19, and classified them further into symptomatic and asymptomatic, we believe that the potential for bias is minimized. Further, we have included both inpatients who are negative as well as asymptomatic positive patients in our ongoing analysis. Most importantly, because the underlying basic science of Raman spectroscopy has been proven to generate a unique spectral signature for each different organism, such as influenza, Ebola, or SARS-CoV-2, in a specific extra-corporeal specimen, we believe the same likely holds true for different organisms examined transcutaneously (5,10).

We are currently working on other biomedical applications for our technology, such as infections, pathology, and cancer diagnosis. We aim to solve existing dilemmas involving intraoperative margin status in cancer surgery, infections (bacterial, viral, or fungal) and pathology with novel devices, such as ours, with further research.

The secondary outcomes analysis from our data is ongoing and will be included as part of future research. As we measured viral load in accordance with the PCR cycle number, we anticipate correlating our device’s test results with the cycle number and other parameters. Using our device to better detect asymptomatic COVID-19 and degree of infectivity is among the goals. We now know that our device can detect COVID-19 among symptomatic patients even more powerfully than the statistics indicate because the study group included both symptomatic as well as asymptomatic individuals in both the PCR negative and positive groups. What the current analysis proves is that with more data, the predictive power increases, and that increasing data will likely have the same effect on the asymptomatic group. Quantifying someone’s immunity, by looking not only at antibody level but also at other immune markers, is another goal. We have collected vaccine information from each patient.

Although a library of Raman Spectra already exists for laboratory-examined infectious agents and many other molecules associated with disease, and is likely not to differ significantly from our transcutaneously-derived library, we hope to add to our own transcutaneously-derived library of infectious agents and molecules. Our study shows that as we added more spectroscopic data to our machine learning algorithm, the predictive power of the device increased. A similar strategy using our device for other pathogens or variants can be used. Using probabilistic models and computational biology, we hope to depict the physical structure of future variants. We could possibly then, using machine learning, determine the Raman signature of future variants. In this way, we will be able to recognize a variant before it is actually sequenced in the lab (21).

As is the case with many hardware-software devices, such as computers, mobile phones, and pulse oximeters, our device will continue to evolve both with its miniaturization and applications.

## Data Availability

All data produced in the present study are available upon reasonable request to the authors.

## Acknowledgement

Rohit Singh, PhD, MIT for his machine-learning expertise.

## References

1. Jegerlehner S, Suter-Riniker F, Jent P et al. Diagnostic accuracy of a SARS-CoV-2 rapid antigen test in real-life clinical settings. Int J Infect Dis. 2021 Aug; 109:118–122

2. Schuit E, Venekamp R, Hooft L et al. Diagnostic accuracy of covid-19 rapid antigen tests with unsupervised self-sampling in people with symptoms in the omicron period: cross sectional study. BMJ 2022;378:e071215

3. Nicolson F, Kircher, MF, Stone, N, et al. Spatially offset Raman spectroscopy for biomedical applications. Chemical Society Reviews; The Royal Society of Chemistry 2021; Issue 1, 2021. DOI/10.1039/DOCS00855A.

4. Andreou C, Kishore SA, Kircher, MF. Surface-Enhance Raman Spectrosopy: A New Modality for Cancer Imaging. J Nucl Med. 2015 Sep;56(9);1925–9.

5. Huang J, Wen J, Zhou M, et al. On Site Detection of SARS-CoV-2 Antigen by Deep Learning-Based Surface-Enhanced Raman Spectroscopy and Its Biochemical Foundations. Anal. Chem. 2021; 93: 9174-9182. Science. Jan 15 2021; 371; 6526:284–288. DOI: 10.1126/science.abd7331

6. Carlomagno C, Bertaziolo D, Gualerzi A, et al. COVID-19 salivary Raman fingerprint: innovative approach for the detection of current and past SARS-CoV-2 infections. Scientific Reports (2021) 11:4943; https://doi.org/10.1038/s41598-021-84565-3

7. Brinati D, Campagner A, Ferrari D, et al. Detection of COVID-19 Infection from Routine Blood Exams with Machine Learning: A Feasibility Study. Journal of Medical Systems; 44; Article number:135 (2020)

8. Aluko OM, Lawal SA, Reuben CS, et al. Understanding the Systemic Effects of COVID-19: Possible Clues to Potential Therapeutic Approaches. DOI: 10.23937/2643-461X/1710057

9. Roberts I, Muela MW, Taylor JM et al. Untargeted metabolomics of COVID-19 patient serum reveals potential prognostic markers of both severity and outcome. Metabolomics.2021 Dec 20;18(1):6

10. Vallejo-Perez MR, Sosa-Herrera JA, Navarro-Contreras HR, et al. Raman Spectroscopy and Machine-Learning for Early Detection of Bacterial Canker of Tomato: The Asymptomatic Disease Condition. Plants (Basel). 2021 Aug; 10(8): 1542. Published online 2021 Jul 28. doi: 10.3390/plants10081542

11. Zhang L, Li C, Peng D, et al. Raman spectroscopy and machine learning for the classification of breast cancers. Spectrochim Acta A Mol Biomol Spectrosc. 2022 Jan 5;264:120300. doi: 10.1016/j.saa.2021.120300. Epub 2021 Aug 21.

12. Ember K, Daoust F, Mahfoud M, et al. Saliva-based detection of COVID-19 infection in a real-world setting using reagent-free Raman spectroscopy and machine learning. Journal of Biomedical Optics; Feb 2022;Vol.27(2).

13. Kang JW, Park YS, Chang H, et al. Direct observation of glucose fingerprint using in vivo Ra man spectroscopy. Sci Adv 2020; Jan 6 (4).

14. Sjoding MW, Dickson RP, Iwashyna TJ, et al. Racial Bias in Pulse Oximetry Measurement. The New England journal of medicine 2020;383(25):2477–78. doi: 10.1056/NEJMc2029240 [published Online First: 2020/12/17]

15. Mosca S, Conti C, Stone N, Matousek P. Spatially offset Raman spectroscopy. Nature Reviews Methods Primers volume 1, Article number:21 (2021).

16. Yeh Y-T, Gulino K, Zhang Y, et al. A Rapid and label-free platform for virus capture and identification from clinical samples. DOI.org/10.1073/pnas.1910113117; December 27, 2019; 117(2).

17. Shipp DW, Rakha EA, Koloydenko AA, et al. Intra-operative spectroscopic assessment of surgical margins during breast conserving surgery. Breast Cancer Res. 2018; 20:69.

18. Zhang L, Wu Y, Zheng B, et al. rapid histology of laryngeal squamous cell carcinoma with deep-learning based stimulated Raman scattering microscopy. Theranostics 2019; 9(9):2541–2554.

19. Lipson J, Bernhardt J, Block U, et al. Requirements for Calibration in Noninvasive Glucose Monitoring by Raman Spectroscopy. Journal of Diabetes Science and Technology; Volume 3, Issue 2, March 2009; 233–241.

20. Kolgiri V, Patil VW. Protein carbonyl content: a novel biomarker for aging in HIV/AIDS patients. Braz J Infect Dis; 2017 Jan-Feb;21(1):35–41

21. Hie B, Zhong ED, Berger B, et al. Learning the language of viral evolution and escape. Science. 2021 Jan 15;371(6526):284–288.doi: 10.1126/science.abd7331.

22. Zhang S, A.M. van der Mee F, Erckens R, et al. Raman spectroscopic detection of interleukin-10 and angiotensin converting enzyme. Journal of the European Optical Society-Rapid Publications volume 17, Article number: 7 (2021)

23. O-Cuevas E, Badillo-Ramirez I, Islas S, et al. Sensitive Raman detection of human recombinant interleukin-6 mediated by DCDR/GERS hybrid platforms. RSC Adv., 2019, 9, 12269

24. Chaudhary N, Nguyet T, Nguyen Q, et al. Discrimination of immune cell activation using Raman micro-spectroscopy in an in-vitr0 and ex-vivo model. Spectrochimica Acta Part A: Molecular and Biomolecular Spectroscopy Volume 248, 5 March 2021, 11911

25. Bergholt M, Hassing S. Quantification of C-Reactive protein in human blood plasma using near-infrared Raman spectroscopy. The Analyst 134(10):2123–7,October 2009

26. Staritzbichler R, Hunold P, Estrela-Lopis I, et al. Raman spectroscopy on blood samples of patients with end-stage liver disease. September 7, 2021,https://doi.org/10.1371/journal.pone.0256045

27. De La O E, Badillo-Ramirez I. Sensitive Raman detection of human recombinant interleukin-6 mediated by DCDR/GERS hybrid platforms. RSC Advances 9(22):12269–12275;April 2019

28. Balan V, Mihai CT, Cojocaru FD et al. Vibrational Spectroscopy Fingerprinting in Medicine: from Molecular to Clinical Practice. Materials 2019, 12(18),2884; https://doi.org/10.3390/ma12182884

